# Complex and High-Risk Percutaneous Intervention Assisted By Extracorporeal Membrane Oxygenation (ECMO)

**DOI:** 10.1101/2023.05.26.23290621

**Authors:** Kifayat Ullah, Liu Bin

## Abstract

**Background:** Despite improvements in PCI techniques and equipment, there are still cases where traditional PCI is insufficient to manage complex and high-risk lesions. Patients with these lesions have increased rates of major adverse cardiac events, including myocardial infarction, cardiogenic shock, and death. In recent years, the use of extracorporeal membrane oxygenation (ECMO) during PCI has emerged as a potential solution to managing complex and high-risk lesions.

**Methods:** This retrospective study included patients who underwent elective complex and high-risk percutaneous coronary interventions with hemodynamic support provided by Veno-Arterial external membrane oxygenation (VA-ECMO) from 2018 to 2022. Complications related to VA-ECMO rates, Complications related to PCI, death, and major cardiovascular, cerebral events (MACCE) during hospitalization and after one-year follow-up were analyzed.

**Results:** This retrospective study overall included 81 patients in which Males (N=60, 74.1%) and females (N=21, 25.9%) having (Average age: 62.74 ±10.807 years) underwent complex and high-risk percutaneous coronary intervention assisted ECMO. The VA-ECMO support was provided for an average of 21.0 hours (With a range of 1-312). Intra-aortic Balloon Pump IABP support was done in 32.1% of patients. The pre-and post-PCI SYNTAX scores of the patients were 39.92 ± (6.4) and 6.04 ± (9.25) respectively. (P <0.001). Most of the patients had triple vessel coronary disease which was the common coronary lesion (47%). Interoperated complications include Cardiac Tamponade (N=1,1.2%), Acute Myocardial Infarction (N=6,7.2%), Cardiogenic Shock (N=2,2.4%), Cardiac Arrest (N=2,2.4%), Arrhythmias malignant in nature which required electro cardioversion (2,2.4%), Ventricular tachycardia (N=1,1.2%), Non-infectious multiple organ failure MODS(N=1,1.2%), Aortic Dissection Type-A (N=1,1.2%). Blood hemoglobin Pre-CHIP assisted VA-ECMO PCI and Post-procedure were 136.17 ± 21.479 g/L and 106.67 ± 19.103 g/L respectively P<0.001). eGFR pre and post-PCI were 67.22 ± 26.85 and 60.09 ± 27.78 respectively (<0.002), Pre and Post PCI EF were 38.69 ± 13.65 and 43.55 ± 13.72 respectively (<0.001), During hospitalization the outcomes for the CHIP assisted by ECMO procedure include Death(N=16,19.8%), Inguinal Hematoma (N=2,2.5%), Bleeding from the punctured site (N=2,2.5%), Peudoaneurysm (N=1,1.2%), Cerebral Infarction(N=1,1.2%), Subarachnoid hemorrhage (N=1,1.2%), No lower limb ischemia, No acute renal injury, Bacteremia, were noted in any of the hospitalization. Patient Hb decline requiring blood transfusion therapy was (N=59, 72.8%). Survival at discharge (Healthy) was (N=65, 80.2%). After one year of post-operation death (N=6, 7.2%)

**Conclusion:** In conclusion, ECMO-assisted high-risk PCI proves to be a safe and effective strategy for complex procedures in patients who are not candidates for Coronary artery bypass grafting (CABG). The use of VA-ECMO resulted in minimal complications and low rates of MACCE during hospitalization and one-year follow-up. Further research is needed to determine the optimal timing for VA-ECMO initiation.

## Introduction

According to the current guidelines coronary artery bypass grafting (CABG) and percutaneous coronary intervention procedures are revascularization elective strategies in complex coronary artery diseases including multiple vessel involvement, bifurcation stenosis, unprotected left main stenosis, and chronic total occlusion CTO [1]. Coronary artery bypass grafting CABG is recommended by guidelines in complex and high-risk coronary artery disease, but for patients who are not suitable candidates for CABG, usage of PCI as revascularization is growing [2,3,4], In patients with complex coronary artery lesions revascularization with PCI or CABG benefits in prognosis [5,6].In these patients, the revascularization proportion is low [7, 8]. Revascularization in complex and high-risk coronary artery lesions is achieved via High-risk (HR-PCI). In such HR-PCI procedures coronary artery dissection, no coronary artery reflow, hemodynamic insatiability, cardiac Tamponade, and sudden cardiac arrest are several complications are related [3]. Yet for interventional cardiologists to operate on, HR-PCI poses a great challenge. In accordance with current available literature on state topic, with mechanical circulatory assistance device during the revascularization can be achieved in complex and high-risk PCI [3, 4, 9]. Mechanical devices can be used as circulatory assistance during HR-PCI include intra-aortic balloon pump (IABP) counter-pulsation, extracorporeal membrane oxygenation (ECMO), Impella (Abiomed, Danvers, MA, US), and Tandem Heart (LivaNova Medical Technology Co., Ltd., Pittsburgh, PA US Co., Ltd., Pittsburgh, PA, United States) [4, 10]. Cardiac arrest or hemodynamic instability can ensue intraoperative HR-PCI, and ECMO can offer prevailing circulatory support and significantly enhance patient prognosis [11]. Though, Veno-arterial (VA)-ECMO support can surge the risk of complications related to ECMO, including an increased risk of infection at the site of intervention, hematoma, destruction of blood cells, and lower limb ischemia are few of the complications associated with ECMO [10]. Still there is a lack of recommended guidelines, as per present, are not as much of clinical data available employing VA-ECMO as mechanical assistance circulatory support in HR-PCI procedure. The aim of the study is set out to examine and analyze the outcomes of preventative use of VA-ECMO during HR-PCI.

## MATERIALS AND METHODS

### Study Population and Design

This single-center retrospective observational study from the second Norman Bethune hospital of Jilin university included 81 patients who underwent elective complex and high-risk percutaneous coronary interventions with hemodynamic support provided by VA-ECMO from 2018 to 2022 and follow up of one-year post-PCI. The patient’s age range was 18 years or older with a diagnosis of complex and high-risk coronary artery disease. VA-ECMO was signposted for the patients having the following criterion I) Having a Left ventricle Ejection Fraction LVEF of ≤30%; (ii) LVEF ≥30% along with the following conditions; (a) LM coronary artery Unprotected, (b) Chronic total occlusions (CTOs) in one or two coronary arteries, in addition, one severe stenosis, (c) Calcified coronary artery lesions requiring rotational grinding and maneuvering for the severity and diagnostics purpose. New onset and older myocardial infarction, and clinical unstable angina pectoris, cardiogenic shock, recent and chronic Heart Failure were among the indicators present in these patients. Intraoperative or Pre-PCI, ECMO establishment was accomplished on each patient, and the Veno-Arterial-ECMO mode was selected. In all patients included in this study common femoral artery and vein were used for the ECMO intubation, as their diameter is 1-2 mm smaller than the inner diameter of the intubated vessel, arterial cannulas at 15–17 (Fr) and venous cannulas at 19–21 (Fr) were selected to avoid complications associated with ECMO. Intubation for VA-ECMO was achieved with guidance of fluoroscopy. Heparin 100 U/kg was an anticoagulation stratagem was used before the Veno-arterial ECMO insertion. The activated clotting time (ACT) during the establishment of VA-ECMO was set at ≥250 sec, then during procedure PCI for 250– 350 seconds. Before PCI and regularly afterward, all patients received 300 mg of aspirin and 180 mg of ticagrelor or 300 mg of clopidogrel PO. Preliminary VA-ECMO blood flow was established at 1.5-2.0 L/min per patient weightiness, and it was later modified in response to a patient’s hemodynamics.

Patients who decline coronary artery bypass graft (CABG) were evaluated by the interventional cardiologist’s team at the study center for HR-PCI assisted by VA-ECMO. Patients with coronary artery disease needed percutaneous coronary intervention (PCI) for revascularization. Raw data for this study collected by accessing patients medical records, inpatients records and follow-ups of the same patients including the baseline demographic and clinical characteristics of patients including age, sex, body mass index, medical history, and pre-procedure medications, intraoperative, and follow up of the patients for major adverse cardiac and cerebrovascular events MACCE events, as hospitalization due to heart failure, stroke, recurring MI, and all-cause mortality. The primary endpoint of this study was the occurrence of major adverse cardiac and cerebrovascular events (MACCE) within the hospital and after the HR-PCI procedure assisted by VA-ECMO. MACCE was defined as (I) composite of death by any of cerebrovascular or myocardial infarction, stroke, and either by re-PCI or CABG-targeted vessel revascularizations. Secondary endpoints included individual components of MACCE, bleeding events, the need for re-hospitalization, and long-term outcomes up to 1 year.

Patient follow-up for 1 year Post-procedure. Clinical assessments and laboratory tests were performed at each follow-up visit to assess for the occurrence of adverse events and to evaluate the long-term outcomes of the procedure. Any adverse event that occurred during the follow-up period was recorded. The procedural details include the type of PCI, and number of stents used. In addition, laboratory data including hemoglobin level, Kidney function test, and liver function test Pre and Post procedure were collected from the in-patient records department. Data regarding ECMO support, including the duration of ECMO support, type of ECMO cannulas, and complications related to ECMO support such as bleeding or thrombosis. The need for additional interventions such as a decompression of LV by intra-aortic balloon pump (IABP) or surgical intervention for ECMO related complication was acquired from the catheterization lab.

Following was considered to be an acute kidney injury (AKI): (i) Rise in serum Creatinine (SCr) of more or equal to 0.3 (mg/dl (≥26.5 μmol/L)) over the course of 48 hours; (ii) a rise in SCr (serum creatinine) ≥1.5 that of the reference value, which known to have or assumed to have happened during the preceding week; and (iii) For six hours the amount of urine < 0.5 mL/Kg/hr [12]. Acute myocardial infarction was defined by the fourth edition of the global myocardial infarction as an upsurge or reduction in blood plasma levels of cardiac troponins that is by at least one time more than the upper limit of the normal range and accompanied by simultaneous clinical evidence of acute myocardial ischemia, such as [13]: (A) Acute myocardial ischemic clinical manifestations, (B) Pathogenesis of the Q wave (novel), (C) Novel noticeable myocardial loss, Segmental wall motion abnormality in LV, (D) Electrocardiogram novel changes of ischemia, (E) Coronary artery angiography imaging examination results, and validation of coronary artery thrombosis were all illustrations of acute myocardial ischemia. Following criterion was used to state coronary artery diseases [14] (i) Micro vasculature dysfunction and/or coronary artery spasm that induces chest-related symptoms in patients who are exposed to stress, exercise or even at rest (Unstable Angina) (ii) ≥50% stenosis of Left main coronary artery and, (iii) ≥70% stenosis in one or more CA. Chronic Total Occlusion (CTO) was stated, to be coronary artery obstruction thru positive thrombolysis having TIMI distal blood flow level 0 and ≥3 months for occlusion. Ipsilateral - collateral vessels or bridging at full occlusion is still taken into account even if distal blood flow TIMI level >0 in an occluded vessel [15]. The SYNTAX score for the patients was calculated online http://syntaxscore2020.com/. For the bleeding incidents, the Bleeding Academic Research Consortium (BARC) criteria were applied [16]. The need for the patient’s informed consent was waived off as it is a retrospective observational study.

### Clinical Outcomes

The outcomes of the study is procedural success, defined as achievement of complete revascularization with a residual stenosis of less than 30%, and in-hospital mortality including the incidence of MACCE events, bleeding events such as hematomas, Peudoaneurysm, Fistula (Atrio-vensous, deep venous thrombosis, acute kidney injury,and or bacteremia. The primary endpoint of this study was the occurrence of major adverse cardiac and cerebrovascular events (MACCE) within the hospital Post PCI. MACCE was defined as (I) composite of death due, cerebrovascular or cardiovascular, stroke, either by PCI or CABG-targeted vessel revascularization. Secondary (Safety) endpoints included individual components of MACCE, bleeding events, need for re-hospitalization, and long-term outcomes up to 1 year.

### Statistical Analyses

Descriptive statistics were used to summarize patient characteristics, procedural details, and outcomes. Continuous variables were presented as mean ± standard deviation those with normal distribution, or median with interquartile range, and categorical variables were presented as frequencies and percentages. Other Statistical tests for normality are Kolmogorov-Smirnov and Shapiro-Wilk. Paired t-test was used for the variables which were relatable and compared their mean of Pre-Op and Post-Op. The test was two-tailed and p-<0.05 statistically significant was set for statistical analyses which were done by IBM SPSS Statistics 27.0 (IBM Corp., Armonk, NY, USA).

## RESULTS

### Baseline Clinical Characteristics

In this retrospective study, total 81 patients were included in which Males (N=60, 74.1%) and females (N=21, 25.9%) having (Average age: 62.74 ±10.807 years) underwent complex and high-risk percutaneous coronary intervention assisted VA-ECMO. The majority of the patients had prior comorbidities. Pre- and post-operatively LVEF was 38.65 ± 13.576 and 43.52 ± 13.640 respectively. All the patients were hemodynamically stable pre-operatively with that of ST-elevated myocardial Infarction (40.7%), non-ST-elevated myocardial infarction (23.5%), and Unstable angina (25.9%). More than half of the patients were of heart failure (50.6%). Patient included in this study was evaluated on basis of New York Heart Association NYHA criteria, most common were NYHA class IV patients (39.5%). All the patients’ went under complex and high-risk PCI assisted by VA-ECMO were elective and rejected the coronary artery bypass grafting CABG. Baseline Clinical Characteristics, comorbidities, and medications before the procedure of patients included in our study are summarized in **Table-1**

**Table 1.** Baseline Clinical characteristics of all the patients included in this study.

| **Parameter** | **ECMO (N=81** |
| --- | --- |
| Age (Years) | 62.74 ± (10.807) |
| Gender-Male | 60 (74.1%) |
| Female | 21 (25.9%) |
| Body Mass Index (Metric Units) | 24.9 ± (3.45) |
| Hypertension | 35 (43.2%) |
| Diabetes | 21 (25.9%) |
| Chronic Kidney Disease | 4 (4.9%) |
| Pulmonary Disease | 6 (7.4%) |
| Atrial fibrillation | 5 (6.2%) |
| Smoking | 24 (29.6%) |
| Hyperlipidemia | 7 (8.6%) |
| Prior Stroke | 6 (7.4%) |
| Prior CABG | 2(2.5%) |
| Prior PCI (Stent restenosis) | 6 (7.4%) |
| Clopidogrel | 66 (81.5%) |
| Ticagrelor | 13 (16.0%) |
| Lung Disease | 19(23.5%) |
| NSAID | 30(37.0%) |
| NYHA Class combined | 2.8 ± (1.1) |
| Class I | 12 (14.8%) |
| Class II | 20 (14.7%) |
| Class III | 17 (21.0%) |
| Class IV | 32(39.5%) |
| **LVEF Pre-Op** | **38.65 ± 13.576** |
| STEMI | 33 (40.7%) |
| NSTEMI | 19(23.5%) |
| Unstable Angina UA | 21 (25.9%) |
| Heart failure (HF) | 41 (50.6%) |
| Refused CABG | 81 (100%) |
| Platelets *10<sup>9</sup>/L | 229.9 ± (79.9) |
| Prothrombin-Time (sec) | 11.86 ± (7.7%) |
| Thrombin-Time (sec) | 14.28± (4.44) |
| Antithrombin (%) | 88.54± (18.70) |
| INR | 1.0 ± (.74) |
| Alanine Aminotransferase (ALT U/L) | 38.75 ± (131.34) |
| Aspartate Aminotransferase<br>AST (U/L) | 42.00 ± (257.8) |
| Albumin (g/L) | 35.89 ± (5.18) |

Data presented as N (%), Means ± SD, and or Medians (Interquartile Q1-Q3). CABG, Coronary Artery bypass surgery; PCI, Percutaneous coronary interventions; NSAID, non-steroid anti-inflammatory drug; NYHA, New York Heart Association.

Intraoperative procedural and angiographic data are summarized in **Table 2**. The pre and post-PCI SYNTAX scores of the patients were 39.92 ± (6.4) and 6.04 ± (9.25) respectively. (P <0.001). Most of the patients had triple vessel coronary disease patients (47%).Maximum number of diseased vessels in single patient was five (5) (N, 1=1.2%). CTO (Chronic total occlusion) lesions was present (41.5%) of the patients per single vessels or in multi-vessels. LAD was the most common coronary disease vessel (95.1%) in patients, followed by RCA and LCX (85.2%) and (79.0%) respectively. LM diseased coronary artery (43.2%). Revascularization was achieved in all the diseased vessels by implantations of (an average of 3.0 (0-6) stents. There was only one patient (1.2%) in which there were no stents deployed because the guide wire did not pass the lesion due to heavy calcification and high tortuosity. In another patient (1.2%) no stent was deployed as there was thrombus aspiration done on that patient successfully, with distal flow TIMI III with no obvious stenosis. In one another patient with stent restenosis (1.2%) had only a single stent implantation, and old stents re-inflated with distal TIMI III flow. 52 patients (64.2%) had ECMO setup intraoperative while 29 (35.8%) had pre-operatively. ECMO weaning time was 21 hours with a range (1-312 hours). Intra-aortic Balloon Pump IABP counter pulsation was used in 32.1% of patients. Need and reason for the counter pulsation for the following; Retaining of contrast agent in coronary sinuses, Due to extended duration of VA-ECMO causes burden left side of the heart, Blood stasis noted in left ventricle on bedside cardiac ultrasound, weaning off VA-ECMO support from patients with poor cardiac function maintained by counter pulsation IABP. Intraoperative complications include Cardiac Tamponade (N=1,1.2%), Arrhythmias malignant required electro cardioversion (2,2.4%), Ventricular tachycardia (N=1,1.2%), MACCE events and other complications during hospital post PCI were Acute Myocardial Infarction (N=6,7.2%), Cardiogenic Shock (N=2,2.4%), Cardiac Arrest (N=2,2.4%),,Non-infectious multiple organ failure MODS(N=1,1.2%), Aortic Dissection Type-A (N=1,1.2%). No patient died during the complex and high-risk PCI procedure.

**TABLE 2.** Procedural & Angiographic parameters of the patients included in the study.

| **Parameter** | **ECMO N (81)** |
| --- | --- |
| Pre-PCI SYNTAX Score | 39.92 ± (6.4) |
| Post-PCI SYNTAX Score | 6.47 ± (9.25) |
| Number of coronary vessels having the disease |  |
| One | 1 (1.2%) |
| Two | 6 (7.4 %) |
| Three | 38 (47 %) |
| Four | 35 (43.2 %) |
| Five | 1 (1.2%) |
| CTO | 34(42.0 %) |
| Location of Lesion CA (Combined) |  |
| Left Anterior Descending | 77 (95.1 %) |
| Left Circumflex | 64 (79.0 %) |
| Right Coronary Artery | 69 (85.2 %) |
| Left Main | 35 (43.2 %) |
| Ramus | 2 (2.46%) |
| OCT | 14 (17.3%) |
| Number of Stents implanted | 3.0 (0-6) |
| No PCI | 1 (1.2 %) |
| Drug Coated Balloon | .35 (0-4) |
| Proglide Use | 81 (100%) |
| IABP counter pulsation | 26 (32.1%) |
| Canulation for distal perfusion | 0(0) |
| Malfunctioning device | 0(0) |
| Non-Invasive Ventilator | 52 (64.2%) |
| **ECMO setup** |  |
| **Intra-Operative** | 52 (64.2%) |
| **Pre-Operative** | 29 (35.8%) |
| ECMO weaning Time (Hours) | 21 (1.0-312) |
| **MACCE in CATH LAB** |  |
| Cardiac Tamponade | 1 (1.2%) |
| Malignant Arrhythmias required electro cardioversion | 2 (2.4%) |
| Ventricular Tachycardia | 1 (1.2%) |
| Death | 0(0) |

Data presented as N (%), Means ± SD, and or Medians (Interquartile Q1-Q3). CA, Coronary artery; PCI, Percutaneous coronary interventions; CTO, Chronic total occlusion; OCT, Optical coherence tomography; IABP, Intra-aortic balloon pump; ECMO, Extracorporeal Membrane oxygenation;

**Table-3** summarized the patients’ blood work, renal function parameters and echocardiography indices pre and post-complex and high-risk VA-ECMO assisted PCI. Blood hemoglobin Pre and Post-procedure were 136.17 ± 21.479 g/L and 106.67 ± 19.103 g/L respectively (P<0.001). Creatinine and blood urea nitrogen were not significantly altered before and after the procedure. Uric acid pre and post-procedure were 435.4 ± 136.5 and 362.9 ± 138.0 (p<0.001) correspondingly. eGFR pre-procedure were 67.22 ± 26.85 and post-procedure minimum 60.09 ± 27.78 (<0.002), Before PCI LVEF was 38.69 ± 13.65 and after PCI were 43.55 ± 13.72 (p<0.001)

**TABLE 3.**
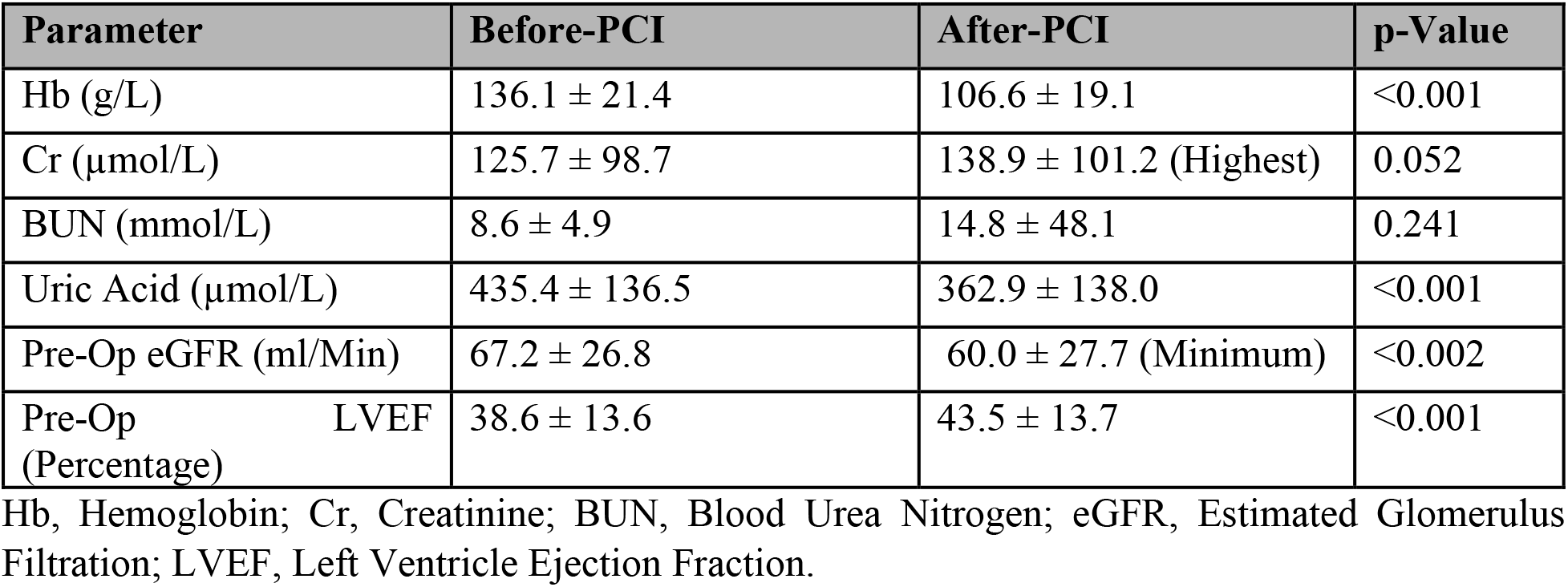
Pre and Post PCI Evaluation Laboratory, Cardiac Indices, and Renal Function parameters

### Clinical Outcomes

**Table-4** Evaluation of patient’s clinical outcomes for CHIP assisted by ECMO procedure include the survival at discharge in which overall procedure was completely done and successfully achieved revascularization without mortality due MACCE or any other complications was in 80.2% (N=65) patients. Death occurs in (N=16, 19.8 % combined all-cause mortality), Re-infarction post procedure in patient stay at hospital occurs in (N=6, 7.5%), Cardiac arrest in 2 (2.5%). Cardiogenic shock (N=2, 2.5%). One patient (1.2%) NSTEMI. Aortic dissection Type-A in one patient (1.2%), Inguinal Hematoma (N=2, 2.5%), Bleeding from the punctured site (N=2, 2.5%) ARC’s-Type-I, Peudoaneurysm (N=1, 1.2%), Cerebral Infarction (N=1, 1.2%), Subarachnoid hemorrhage (N=1, 1.2%), Patients with electrolytes imbalance, contrasts clearance and decompression on left ventricle were treated with continuous renal replacement therapy (N=26, 32.1%), No lower limb ischemia, no acute renal injury, Bacteremia, during the stay at the hospital post procedure. Patient Hb decline requiring blood transfusion therapy in (N=59, 72.8%).

**TABLE 4.**
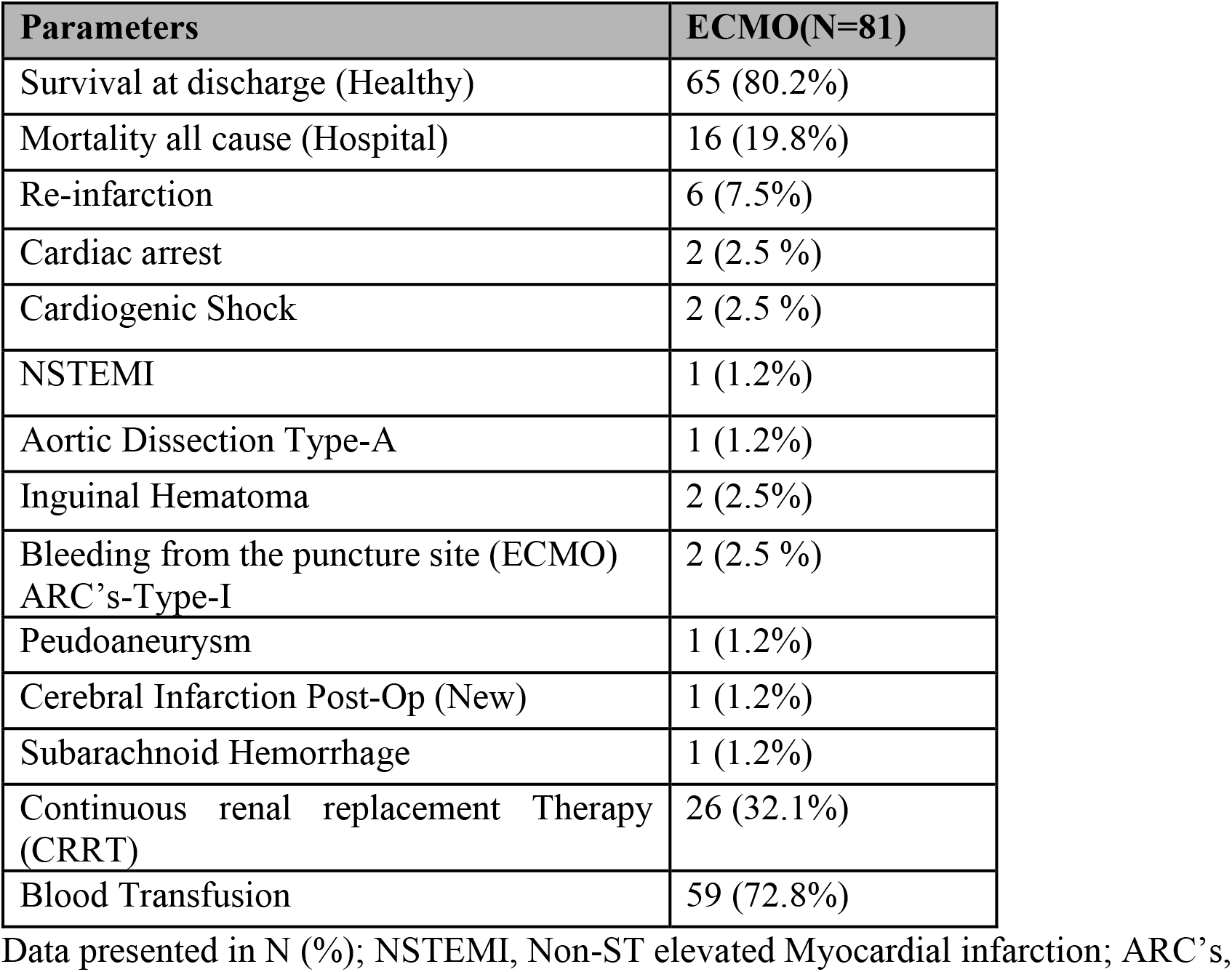
Clinical outcomes of the study during the stay at the hospital

**Table-5** summarized the occurrence of MACCE, other major complications, and all-cause mortality over one-year follow-up after the complex and high-risk PCI assisted by VA-ECMO. After one year of follow-up (N=29, 35.8 %) were healthy without any complications and MACCE events with a time duration of 7.02 months with a range (0-34 months), zero (0) for the patients who completely lost after the procedure and thirty-four (34) is the longest follow up duration. Out of a total of 27 (33.3%) patients lost to follow-up in which 5 patients (6.2%) follow up for one (1) month and Twenty-two (N=22,27.1%) post-PCI never follow-up. Six (6, 7.5%) died over the course of one year of follow in which one patient died of ventricular fibrillation after discharge, Aortic stenosis occurred in one patient after one month of high-risk PCI, one patient died of terminal illness, Recurrent acute myocardial infarction (stent restenosis) occurred in one patient six months post-PCI, one patient died of acute heart failure after 28-days, one patient died of non-infectious multiple organ dysfunction syndrome (MODS). Angina occurred in two (N=2, 2.5%) seven and sixteen months postoperative respectively, both patients received appropriate treatment and are currently in healthy condition.

**TABLE 5.** Outcomes and MACCE over the 1-year follow up

| Parameters | ECMO (N=81) |
| --- | --- |
| Healthy | 29 (35.8%) |
| Time Duration (Months) | 7.02 ± (10.0) |
| Lost to follow-up (Combined) | 27(33.3%) |
| Died (Combined) | 6 (7.5%) |
| Ventricular Fibrillation | 1 (1.2%) |
| Aortic Stenosis | 1 (1.2%) |
| Terminal illness | 1 (1.2%) |
| Recurrent Acute MI | 1 (1.2%) |
| Acute Heart failure | 1 (1.2%) |
| Non-Infectious MODS | 1 (1.2%) |
| Angina (Improved) | 2 (2.5%) |

Data presented in mean ± standard deviation, N (%); MI, Myocardial Infarction; MODS, multiple organ dysfunction syndrome;

## Discussion

Revascularization in patients with complex and high-risk coronary artery disease has long posed challenge for interventional cardiologists due to inherent risks and potential complications associated with the procedure. These patients are characterized by significant coronary artery disease and related complications, intraoperative-ly they are prone to hemodynamic instability. CABG a preferred choice of treatment in patients SYNTAX score ≥23 having left main coronary artery disease, or in patients SYNTAX score ≥23 with triple vessel disease with or without diabetes. Revascularization with percutaneous coronary artery intervention is appropriate management in one or two coronary arteries lesions in elective revascularization strategy in complex and high-risk coronary arteries with or without stenosis of left anterior descending. [1] The findings from analyzing 81 patients in a study indicate that the utilization of VA-ECMO (veno-arterial extracorporeal membrane oxygenation) in elective high-risk percutaneous coronary intervention (HR-PCI) is safe and viable. The study reported low rates of mortality and complications. Cases involves patients with intricate and high-risk coronary artery disease, selective HR-PCI assisted by VA-ECMO could be considered as an alternative approach to coronary artery bypass grafting (CABG) for elective complex and high risk, or refusal from cardiac surgery. This study outcomes aligns with similar studies conducted at other single-center facilities. [17-23] Patients with complex and high-risk coronary artery disease commonly demonstrate three key clinical characteristics. First, presence of severe coronary artery disease, which includes conditions such as multi-vessel disease or involves unprotected left main trunk, chronic obstructive disease with or without calcification, and subsequent development of complications. Second, involves the coexistence of morbidities such as heart failure, diabetes, a history of previous coronary artery bypass grafting (CABG) or PCI (Stent restenosis), and advanced age. Third, manifestation of hemodynamic changes, which may encompass hemodynamic instability, shock, or severe left ventricular dysfunction [3, 4] In SYNTAX trials, results from ten years post-revascularization all-cause mortality in PCI and CABG, no difference amongst both [2]. In this study HR-PCI triple vessels revascularization in CABG patients was successful but they were very limited in comparison to others in terms longer survival duration and no MACCE events post HR-PCI, Bai et al reported in their study, they formed a subgroup of patients underwent ECMO in elective HR-PCI that triple vessel in CABG patient survival rate was higher as compared to other PCI procedure but in left main stenosis PCI and CABG were same survival rate [21]. Revascularization stratagem in left main or triple vessel coronary artery disease patients were evaluated by cardiac surgeons/interventional cardiologists [2]. CABG or PCI revascularization rate in complex and high-risk coronary artery disease is low. In one group of a study ‘Global Registry for Acute Coronary Events Score, recruited 4,414 patients of NSTEMI (Non-ST elevated myocardial infarction) and divide them into low-risk, medium risk, and high-risk patients. In their study, they found revascularizations in high-risks group was significantly lesser compared to low and medium-risk group. Though, in such complex and high-risk coronary artery disease both CABG and PCI is progressively growing over time [7]. Another observational study evaluated revascularization in multi vessels coronary artery lesions and non-ST elevated myocardial infarction (NSTEMI) with comorbidity diabetes, including 29,769 patients. Their findings suggested of all patients who underwent revascularizations within the span of six years, half of patient went for PCI and one-third for CABG, and the proportion of total revascularization increased. Patients underwent CABG, proportion remains same but increase revascularization with PCI progressively. Revascularization with PCI or CABG in complex and high-risk coronary artery disease enhance prognosis as proposed [5, 6].

Nevertheless, high-risk percutaneous coronary intervention (HR-PCI) presents several challenges in its implementation [3, 9]. Firstly, research data are scarce due to insufficient rates of revascularization and a lack of objective and reliable evidence supporting an optimal strategy for revascularization. Secondly, interventional physicians may have underestimate potential benefits of revascularization in this specific patient population. Thirdly, performing revascularization procedures in complex and high-risk coronary artery disease patients can be challenging, as intraoperative procedures and complications may significantly affect hemodynamic parameters. Lastly, operators must possess expertise in various techniques such as fractional flow reserve, intravascular ultrasound, and optical coherence tomography for better guidance. Consequently, a substantial sum of interventional physicians may lack the essential proficiency mandatory for such procedures in which patients. An increase in clinical evidence of using mechanical assistance devices for the left heart, indicates efficacy of left heart assist devices for providing circulatory support during high-risk percutaneous coronary intervention (HR-PCI). IABP is an older mechanical assistance circulatory device, findings from IABP-SHOCK II trial, a study involving mechanical circulatory assistance devices for hemodynamic supports, demonstrated the ineffectiveness of the intra-aortic balloon pump (IABP) in patients experiencing circulatory failure when used alone [24]. Al-Khadra et al. conducted an assessment of non-emergency percutaneous coronary intervention (PCI) in patients without cardiogenic shock and acute myocardial infarction, utilizing a percutaneous ventricular assist device (PVAD) and intra-aortic balloon pump (IABP) support, respectively. In their findings, they stated mortality rates were lower in that PCI assisted by PAVD than in PCI assisted by the IABP used solely [25]. The use of ECMO (extracorporeal membrane oxygenation) can provide robust mechanical circulatory support for hemodynamic management during elective high-risk percutaneous coronary intervention (HR-PCI) [10, 11]. In context of complex and high-risk PCI, the utilization of VA-ECMO necessitates the comprehensive evaluation of both the patient’s cardiac functional condition and the extent of coronary artery disease by the interventional cardiologist. This evaluation is crucial due to the latent occurrence of significant hemodynamic instability during PCI in patients with severe coronary artery disease. Therefore, for the conclusiveness in this study we includes patients with LVEF ≥15 %. Patients with left main unprotected, dual CTO with one more unembellished coronary artery stricture in which there is need aimed at maneuvering for disease severity or rotational atherectomy required in complex and high-risk PCI needed VA-ECMO support, for that reason we included a vast range (LVEF=15%-LVEF=70%) patients in our study. All patients did well at the end of the procedure without intraoperative mortality or MACCE events leading to mortality. There were four cases in which MACCE occurs in two patients with VF, a VT and one cardiac tamponade ensues but were resolved immediately with electro cardioversion and medical support immediately and appropriately. Re-infarction occurs in (N=6,7.5%) patients, in two patients (2.5%) cardiac arrest occurs which leads to death without any treatment option not attributable to HR-PCI or ECMO-related complication, due to the stent’s thrombosis and restenosis, more the comorbidities, late the interventions,worst ending. NSTEMI was noted in one patient (1.2%) with a previously discussed attributable cause as discussed earlier because these patients did very well in the procedure and ECMO was weaned off successfully after that MACCE events happened. In one patient (1.2%) Type-A aortic dissection occurs which blocks RCA leading to acute myocardial infarctions, was not the complications of HR-PCI VA-ECMO but sudden spike in blood pressure. Peudoaneurysm (N=1,1.2%) in one patient, repaired surgically without leading to in-hospital mortality, Cerebral infarction was noted in one patient (1.2%) lacking any serious complication or mortality countered by anticoagulation strategy in the patient, and subarachnoid hemorrhage occurs in (N=1,1.2%) patient, Both cerebral infarction and subarachnoid hemorrhage occurs post-procedure due to prolong durations of ECMO in these two patients which were promptly diagnosis and treated appropriately without causing any mortality in patients were healthy afterward. VA-ECMO mechanical circulatory support in severe coronary artery disease and more prospective and large-sample randomized studies are required. Several studies conducted at single centers, with small sample sizes, have examined the use of VA-ECMO as a mechanical circulatory support strategy for HR-PCI. The findings from their studies indicates that VA-ECMO is safe and effective. Furthermore, elective HR-PCI supported by VA-ECMO proves to be a viable alternative for patients who are not eligible for CABG or are deemed high-risk, offering favorable short-term and long-term prognoses. At present, existing clinical evidence regarding HR-PCI assisted by VA-ECMO is inadequate, necessitating further validation through randomized controlled trials. Successful implementation approach relies on the expertise of specialized teams comprising experienced ECMO and cardiac interventional specialists. ECMO, in comparison to alternative percutaneous mechanical support devices, possess greater operational challenges, clinical advancement and patient outcomes have been hindered by associated complications.[26] Competence in the identification, appropriate on time treatment, and management of ECMO-related complications is largely based on capability of the team in the diagnosis, treatment, and nursing care of ECMO patients. Studies have demonstrated that ECMO centers that manage the workload of more than 20 critically ill patients each year can maintain the expertise needed in ECMO treatment. [27] In addition, centers that specialize in ECMO care for adults, which treat more than 30 cases per year, have significantly lower mortality rates than centers that manage fewer than six cases per year. [28]. In center for this study expert for the ECMO are in accordance with the criterion and have excellent skills set, less VA-ECMO related complications in patients of complex and high-risk coronary artery PCI. The main complications of VA-ECMO in our study are inguinal hematoma, bleeding from the puncture site, elevated pressure on the left side of the heart (LV) counter by the IABP support successfully, blood loss in the VA-ECMO external circuit, there was no deep venous thrombosis, lower limb ischemia, and most importantly no infection or bacteremia was noted in any of the patient included in those with prolonged duration of ECMO in our study. For VA-ECMO intubation, all patients included in this study femoral artery and vein evaluated under the guidance of fluoroscopy by keeping cannula diameter lesser about 1-2 mm that of intubated artery and vein to avoid ipsilateral cannula and lower limb ischemia and thrombosis or DVT. [29]. Based on the experience from our study cannula was intubated under the guidance of fluoroscopy to evade arterio-venous fistula complications related to ultrasound-guided cannulation. Choice of vascular access depends on the patient’s anatomy, comorbidities, and the experience of the interventional team. In addition, the management of anticoagulation during ECMO-assisted PCI is another important consideration. Anticoagulants are essential to prevent thromboembolic events during ECMO-assisted PCI, but it must be balanced against the risk of bleeding. The optimal anticoagulation strategy depends on the patient’s comorbidities, the type of ECMO used, and the individual patient’s response to anticoagulation. In this study, two patients (2.5%) develop a hematoma and two patients (2.5%) have bleeding from the intubation site of cannulation out of a total of 81 patients which were treated appropriately without causing mortality. In (N=59) patients (72.8%) hemoglobin Hb level drops requiring blood transfusion therapy were done primarily due to loss of blood in the VA-ECMO external circuit. In this study left ventricle overload due to prolong use of VA-ECMO support in some was decompressed with counter-pulsation with IABP in (N=26, 32.1%) patients remaining were VA-ECMO without IABP support or after the weaning of ECMO support. Impella (Abiomed) and intra-aortic balloon pump (IABP) are commonly used as primary devices for left ventricular unloading during VA-ECMO. Additional strategies include opening an atrial septum, surgical drainage of the left ventricular apex, use of positive inotropes, use of diuretics, and use of continuous renal replacement therapy. These approaches collectively contribute to the management of left ventricular overload in the setting of VA-ECMO. [10, 11] Ventricular decompression with any of the unloading devices is better and reduced mortality in VA-ECMO-supported patients than in no loading. [30]. Among patients with cardiogenic shock undergoing VA-ECMO, no substantial disparity in hemodynamics parameters was observed by comparing effectiveness of IABP and Impella (Abiomed) for reducing left ventricular afterload. Nevertheless, concomitant utilization of IABP with ECMO may potentially contribute to a reduction in the death rate and an enhanced 180-day survival proportion reported [31]. Amongst left ventricular decompression devices used in VA-ECMO, IABP holds prominence due to its ease of percutaneous bedside implantation and straightforward operability. Combining IABP with VA-ECMO in patients experiencing cardiogenic shock yields notable benefits, including a substantial reduction in all-cause mortality rates during hospitalization and at day-28. Moreover, this approach aids in the successful weaning of patients from ECMO support,

[32] in this study this approach was used in poor left ventricle function. Additional investigation is warranted to determine the optimal timing of IABP implementation as a left ventricular decompression stratagem in the context of selective complex and HR-PCI supported by VA-ECMO.

According to ELSO (Extracorporeal Life Support Organization) between 2014 and 2018, the infection rate midst patients undergoing VA-ECMO was documented at 7.6% [26]. Bacteremia and sepsis are frequently observed as common complications associated with VA-ECMO infections. The incidence of infection tends to rise progressively as the duration of ECMO support extends. Notably, over 53% of patients experiencing infection-related complications encounter them within succeeding two weeks of post-ECMO intubation. [28] In this study, ECMO-related infection during or post-procedure no single case was reported of ECMO-related infection complications, This Is because ECMO intubation in our center is more than 30 per year and less time for ECMO support in total duration is less. The total VA-ECMO support in our study was an average was 21 hours (1-312) with one hour being the minimum and 312 hours being the maximum duration. Published studies suggest that ECMO should be weaned off as early as possible to avoid infection because the longer the duration for the ECMO increased the chances of infection [33]. In our study patients were in more serious state of condition pre-HR-PCI, with unprotected left main, severe coronary artery lesions with and without calcification, multi vessels disease, and depressed organ function such as advanced heart failure, atrial fibrillations, and chronic kidney disease patients, prior stroke were included. With these clinically noticeable characteristics, mechanical assistance circulatory device were given feasible opportunity to step in and assist in complex and high-risk coronary intervention. Acute renal injury (ARF) is prognosticative issue of ECMO. Studies have reported rate of severe ARF is approximately 45% in patients on ECMO requires renal replacement rehabilitation.[35].There are several factors attribute to ARF such as damaged blood cells by ECMO, Inflammatory response, ischemic reperfusion injury, Though ECMO can relieve AKI concomitantly increases the risk of ARF.[36].In 2660 single center study patients of CAD (coronary artery disease) were divided into two groups complex (1532) and noncomplex (1128), their study reported no difference contrast related AKI, no increase in contrast associated ARF in complex group[37]. It is believed ARF is not related with contrast associated. In this study Creatinine (Cr) and Blood Urea Nitrogen (BUN) were not significant pre- and post-operation. There was not patient with acute kidney injury relates to low ARF rate. It is probably due to smaller group of patients and short duration of ECMO support. eGFR and uric acid were significantly changed before and after the procedure but were the same in patients who survived and or dead. Cr, BUN, levels indicate that the more severity of the lesion worse the results. Average survival duration post-procedure was 7.02 months, and the prolonged case follow-up of the patient is 34 months. Which is the prolonged timespan in this study and not present in previous ones. Previous studies indicate that LVEF in complex and high risk improves after the HR-PCI procedure and has fewer mortalities and hospital visits almost half of those with conventional medication therapy patients without complex and high-risk PCI procedures. [34] In comparison with our study, those who received interventional LVEF improves hence cardiac perfusion as compared to those with conventional therapy.

Thou this study gives good clinical results but still, there are certain limitations on the basis of which we cannot generalize it to the rest of the population. Firstly, there was a single-center retrospective study, Secondly, the sample size was small no randomization and no control group.

## Conclusion

ECMO-assisted high-risk PCI is a valuable tool to enhance the safety and efficacy of complex PCI procedures and observing different statistical test elective complex and high-risk PCI assisted by VA-ECMO as mechanical hemodynamic support is a safe and viable option for those patients who refuses CABG or got rejected. VA-ECMO-related complications and MACCE events within hospitalization and after one year of follow-up post-operatively are very low. The Optimum time to introduce the VA-ECMO needs further validation.

## Data Availability

All the data will be shared upon the request

## Abbreviations

VA-ECMO: Veno-arterial Extracorporeal membrane oxygenation
IABP: Intra-aortic balloon pump
CTO: chronic total occlusion
OCT: optical coherence tomography
SD: Standard deviation
CA: Coronary artery
eGFR: estimated glomerulus filtration rate
PCI: percutaneous coronary interventions
LVEF: Left ventricle ejection fraction
CHIP: Complex and High-risk interventional procedure
CABG: Coronary artery Bypass Grafting;

## ETICS STATEMENT

Is this study, direct informed consent was waived off. But before every HR-PCI procedure informed consent was taken from the patients itself or from next of kin. Human participant in this study were reviewed, sanctioned by committee for ethics of the second affiliated Norman Bethune Hospital of Jilin University.

